# Using Natural Language Processing of Clinical Notes to Supplement Structured Electronic Health Record Data for Phenotyping Smoking and Obesity in a Healthcare System

**DOI:** 10.64898/2026.01.18.26344356

**Authors:** Jie Yang, Bowen Gu, Haritha Pillai, Joyce Lii, David Cronkite, Keith A. Marsolo, Rishi J Desai

## Abstract

**Purpose:** Studies based on electronic health records (EHR) often rely on structured data, which may incompletely capture important clinical phenotypes in EHR notes. The purpose of this study was to assess two natural language processing (NLP) tools to extract phenotypes from unstructured EHR notes, and to evaluate the added value of integrating NLP-derived phenotypes with structured EHR data at a health system scale.

**Methods:** This retrospective study is based on inpatient and outpatient EHR data from the Mass General Brigham healthcare system between January 1, 2019 and December 31, 2020. Two established rule-based NLP tools were applied to extract smoking and obesity information from 19,215,303 clinical notes of 503,025 patients. NLP performance was evaluated through manual review of stratified samples. Phenotype prevalence was estimated using structured EHR data alone and compared with prevalence estimates obtained by supplementing structured data with NLP-derived features.

**Results:** Both NLP tools exhibited high performance, with both accuracy and F1 score of 0.99 for smoking, and 0.92 and 0.91 for obesity, respectively. The combination of NLP and structured data identified 220,714 patients (43.88%) with smoking, compared with 170,396 patients (33.87%) identified using structured data alone, representing a 29.5% relative increase. For obesity, NLP identified 121,360 patients (24.13%) from EHR notes, and 169,905 patients (33.78%) were documented in structured data; inclusion of NLP-derived features contributed additional 32,823 patients, corresponding to a 19.3% relative increase.

**Conclusion:** NLP-derived phenotypes from unstructured EHR notes substantially improve patient identification for both smoking and obesity compared with structured EHR data alone at scale.

## 1. Purpose

Electronic health record (EHR) data capture extensive health information generated in real-world clinical scenarios and have therefore been widely used in epidemiologic research and regulatory applications, including post-marketing surveillance.^1,2^ Most existing studies rely primarily on structured EHR data, such as diagnosis, procedure, medication codes and laboratory test.^3,4^ However, structured data often suffer from missing or incomplete entries, which can lead to misclassification, biased estimates, and reduced validity of downstream analyses.^5^

With more clinical detail, unstructured EHR notes can further supplement structured data by capturing patient and disease information, such as symptoms, disease severity and social determinants that are not routinely encoded in structured EHR fields. Such information from unstructured EHR notes can be automatically extracted into structured formats using natural language processing (NLP).^6,7^ However, the performance of NLP-extracted phenotypes (defined here as clinically meaningful patient characteristics) has not been consistently validated in large-scale, real-world healthcare systems. In addition, differences in phenotype prevalence derived from structured EHR fields versus EHR notes and their combination have not been systematically evaluated, it remains unclear to what extent integrating NLP-extracted features with structured EHR data can meaningfully improve phenotype capture at the population level.

This study aimed to validate two NLP-based phenotype extraction tools for smoking and obesity, two common and clinically important risk factors frequently documented in unstructured EHR notes, and to assess the added value of incorporating NLP-derived phenotypes from unstructured EHR notes at a health system–wide scale.^8^ Specifically, we manually evaluated the performance of two EHR-specific NLP tools in extracting smoking and obesity phenotypes. We then compared phenotype prevalence estimated using structured EHR data alone with estimates obtained by supplementing structured data with NLP-derived features, leveraging data from more than 500,000 patients and over 19 million EHR notes from Massachusetts General Brigham healthcare system.

## 2. Methods

This study includes both inpatient and outpatient encounters from Massachusetts General Hospital and Brigham and Women’s Hospital, the two main hospitals within the Mass General Brigham (MGB) healthcare system, the largest healthcare system in Massachusetts, during a two-year period (January 1, 2019 to December 31, 2020). The study sample was derived from a standing data infrastructure of MGB EHRs linked with Medicare or Medicaid claims data, which has been created as a part of the FDA Sentinel initiative.^8^ The final dataset comprised 19,215,303 EHR notes from 503,025 unique patients.

For smoking information extraction, we used the Konsepy pipeline^9^ following the implementation guidelines developed by the team at Kaiser Permanente Washington (KPWA). The tool takes notes as input and extract status with: *A-active, N-negative, H-history and U-unknown*. For obesity information extraction, we used cTAKES^10^ (Clinical Text Analysis and Knowledge Extraction System), a popular open-source NLP system developed by the Mayo Clinic, to extract clinical concepts from EHR notes. To build obesity-related concepts, we queried “obesity” in the UMLS Metathesaurus Browser (https://uts.nlm.nih.gov/uts/umls/home), filtered results by the semantic group “Disorders,” and manually curated a list of 88 relevant CUIs for concept extraction. (list in **supplementary eTable 2**). The cTAKES extracts status from EHR notes with five statuses: *A-active, N-negative, H-history, X-non-patient, and U-unknown*. For both smoking and obesity, NLP outputs for each patient were aggregated into binary “positive” and “negative” groups, as detailed in **Supplementary eMethods**. This aggregation was used to align the NLP-derived phenotypes with the structured EHR comparator based on current or historical evidence of the phenotype. For the evaluation of the NLP performance, we manually reviewed 200 sampled cases for smoking and 250 for obesity, using a stratified sampling strategy. Detailed methods are provided in the **Supplementary eMethods**.

The prevalence of structured EHR labels for smoking and obesity was calculated using the tobacco field and height/weight values, respectively. The tobacco field was available for all patients and categorized as “never,” “current user,” or “quit/former user.” Patients with any tobacco record of “current user” or “quit/former user” during the study period were classified as “positive”. For obesity, BMI was calculated from height and weight in structure EHR records, and patients with any BMI > 30 recorded before December 31, 2020, were classified as “positive”. We further calculated prevalence from NLP outputs and quantified prevalence change when using structured data alone versus when including NLP-extracted features.

## 3. Results

The study included 503,025 patients. Of these, 41.61% were male and 73.50% were White. The median age was 68 years (IQR, 47 to 76) as of December 31, 2020. Detailed demographic characteristics are summarized in **supplementary eTable 1**. Manual review showed that both NLP tools performed well in identifying phenotypes from EHR notes. Specifically, the smoking NLP tool achieved an overall accuracy of 99.50% and an F1 score of 99.34%, and the obesity NLP tool achieved an overall accuracy of 91.60% and an F1 score of 91.26% (**Table 1**). In addition, the negative predictive values were 100% and 90.00% for smoking and obesity, respectively.

**Table 1:** Performance characteristics of the natural language processing (NLP) pipelines for smoking and obesity against manual review.

| Class | Metric (%) | Smoking | Obesity |
| --- | --- | --- | --- |
| Positive | Precision | 99.33 | 92.67 |
|  | Recall | 100 | 93.29 |
|  | F1-score | 99.67 | 92.98 |
| Negative | Precision | 100 | 90.00 |
|  | Recall | 98.04 | 89.11 |
|  | F1-score | 99.01 | 89.55 |
| Overall | Accuracy | 99.50 | 91.60 |
|  | F1-score | 99.34 | 91.26 |

\* Precision corresponds to positive predictive value (PPV), recall corresponds to sensitivity, and F1-score is the harmonic mean of precision and recall.

For smoking, 207,476 patients (41.25%) were identified as positive smokers through unstructured notes, compared to 170,396 patients (33.87%) identified from structured data. Combining patients identified from both structured data and unstructured notes resulted in 220,714 patients classified as smokers (43.88%). Using the structured count as the reference, the NLP method contributed an additional 50,318 patients, representing a 29.5% relative increase (**Figure 1**). For obesity, 121,360 patients (24.13%) were identified through unstructured notes, while 169,905 patients (33.78%) were identified from structured data. The NLP method contributed an additional 32,823 patients, corresponding to a 19.3% relative increase. Among manually validated cases, NLP-only additional cases had PPVs of 93.33% for smoking (15/200 validation cases) and 75.00% for obesity (24/250 validation cases). Compared with patients identified through structured data, NLP-only additional patients were younger for both smoking and obesity phenotypes, with median ages of 66 versus 71 years for smoking and 63 versus 68 years for obesity, respectively (Supplementary eTable 2).

**Figure 1:**
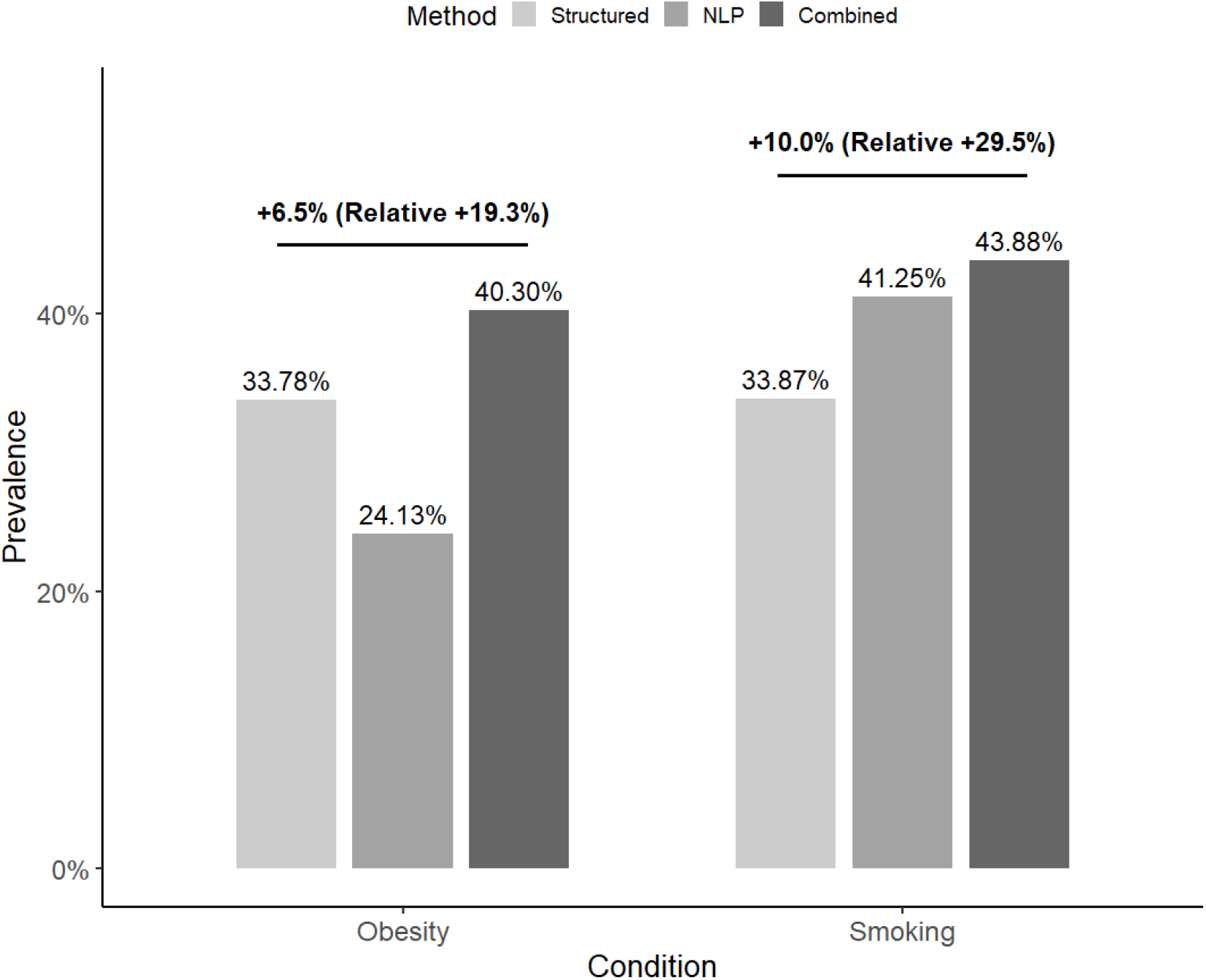
Estimated prevalence of obesity and smoking from electronic health records, Mass General Brigham 2019-2020 (n=503,025) ^*^Footnote: The labels on the bars indicate increase in prevalence estimates when using both structured data and natural language processing (NLP) outputs compared to using structured data alone

## 4. Conclusions

In this study, we validated two NLP phenotype tools for smoking and obesity using more than 500,000 patients and over 19 million EHR notes from a real-world healthcare system. Manual review results demonstrated that both NLP tools achieved high performance, indicating their effectiveness and reliability for extraction of smoking and obesity patients from large-scale unstructured EHR text.

Integrating NLP-derived phenotypes with structured EHR data has shown substantial improvement in phenotype prevalence compared with structured data alone. For both smoking and obesity, NLP identified a meaningful number of additional patients not captured through structured fields, demonstrating under-coding issue in structured EHR elements. In addition, for phenotypes that are not routinely or consistently captured in structured EHR fields, unstructured clinical notes may serve as the primary, and in some cases the only, source of relevant information, making the incorporation of narrative data even more important for accurate population-level analyses.

Both NLP tools evaluated in this study are rule-based approaches, which are well suited for large-scale deployment due to their computational efficiency and interpretability. Both smoking and obesity are relatively straightforward phenotypes that can be reliably identified using rule-based methods, making them appropriate use cases for health system–wide implementation. Although these rule-based approaches enabled efficient processing on millions of clinical notes, they may face challenges in identifying complex phenotypes or those requiring deeper semantic understanding of EHR notes. More advanced NLP methods, such as large language models (LLMs), hold substantial potential for extracting richer, more complex and fine-grained clinical information with potentially better performance.^11-13^ However, they may require significantly greater computational resources and may be less scalable, less interpretable, and more difficult to adapt or implement consistently across large-scale EHR analyses. One potential hybrid approach is to use LLMs to annotate training data for scalable statistical or smaller neural models,^14^ which can then be applied to large-scale extraction.

To our knowledge, this is the first large-scale study to systematically assess and compare phenotyping from both structured and unstructured EHR data using NLP at the level of an entire health system. Several limitations exist in this study. First, we evaluated two separate NLP tools for the two phenotypes, rather than a unified modeling framework. This limits direct methodological comparison between phenotypes. Second, the NLP-derived phenotypes were classified as binary outcomes and did not distinguish current from historical status, which facilitated comparison with structured EHR definitions but may have reduced temporal granularity. Structured and NLP-derived phenotypes may also be imperfectly temporally aligned, particularly for obesity, because structured BMI values could occur before the study period whereas NLP was limited to study-period notes. Third, manual review validation was limited in sample size because of resource constraints; further fine-grained stratification (e.g. by note type and population characteristics) would better characterize NLP performance. Finally, both smoking and obesity phenotypes examined in this study are relatively high-prevalence conditions.

The performance and added value of NLP-based phenotyping for different or low-prevalence phenotypes, such as rare diseases or infrequently documented symptoms, remain to be evaluated.

## 5. Plain Language Summary

Electronic health records contain large amounts of health information in real-world clinical scenarios, but much of this information is documented in free-text clinical notes rather than standardized data fields. In this study, we evaluated two natural language processing tools to identify smoking and obesity phenotypes from EHR notes across a large healthcare system. Using data from more than 500,000 patients and 19 million clinical notes, we found that these tools performed well and identified substantially more patients with smoking or obesity than were captured using structured records alone. These findings show that relying only on structured data can miss important health information and highlight the value of using clinical notes to improve the completeness of health information at the population level.

## Data Availability

Data is not available to the public.

## Acknowledgement

The authors acknowledge support from the US Food and Drug Administration (FDA) through Master Agreement 75F40119D10037 in creating the data infrastructure that was used for this study.

## Ethics Statement

This study was approved by the Mass General Brigham Institutional Review Board.

