## Supplementary for "Using Natural Language Processing of Clinical Notes to Supplement Structured Electronic Health Record Data for Phenotyping Smoking and Obesity in a Healthcare System" (from another version of this preprint: version 1, 2026-01-21)

**eMethods:**

1. **Settings of NLP Tools:**

For smoking, we used the Konsepy pipeline (<https://github.com/kpwhri/fe5_konsepy>) to extract smoking-related information, following the implementation guidelines developed by the team at Kaiser Permanente Washington (KPWA). The tool takes notes as input and extract, status with: *A-active, N-negative, H-history and U-unknown*.

For obesity, we used cTAKES (Clinical Text Analysis and Knowledge Extraction System), a popular open-source NLP system developed by the Mayo Clinic, to extract clinical concepts from EHR notes. To extract obesity-related concepts, we queried “obesity” in the UMLS Metathesaurus Browser (<https://uts.nlm.nih.gov/uts/umls/home>), filtered results by the semantic group “Disorders,” and manually curated a list of 88 relevant CUIs for concept extraction. (list in **supplementary eTable 2**). the NLP tool extract status from EHR notes with five statuses: *A-active, N-negative, H-history, X-non-patient, and U-unknown*.

2. **Phenotype Aggregation for Patient Level**

For both smoking and obesity, patients were classified as “positive” if any of their notes contained mentions of active (*feature_status = A*) or historical (*feature_status =H*) status based on the output of the NLP tools during the study period. Patients with no qualifying entries were assigned the “negative” label.

3. **Sampling Strategy for Manual Review**

Manual chart review was conducted using a stratified sampling strategy to evaluate the performance of NLP-extracted phenotype statuses. For each phenotype, we randomly selected 100 notes labeled with an active status (*feature_status = A*) and 50 notes each from *history (H)*, *negated (N)*, and *non-patient (X)* categories, where applicable. For smoking, the applicable categories were A, N, and H, resulting in a total of 200 reviewed notes. For obesity, the applicable categories were A, N, H, and X, resulting in a total of 250 reviewed notes. All reviews were conducted by a study author (HP) with medical training and master’s degrees in public health.

**eTable 1: Characteries of study population**

| **Data source** | Mass General Brigham |
| --- | --- |
| **Data years** | 2019/1/1-2020/12/31 |
| **Population size** | 503,025 |
| **Basic demographics** |  |
| **Age groups** |  |
| **0-4 yrs** | 1367 (0.27%) |
| **5-9 yrs** | 4602 (0.91%) |
| **10-14 yrs** | 7616 (1.51%) |
| **15-18 yrs** | 7847 (1.56%) |
| **19-21 yrs** | 7223 (1.44%) |
| **22-44 yrs** | 85539 (17.00%) |
| **45-64 yrs** | 85144 (16.93%) |
| **65-74 yrs** | 138154 (27.46%) |
| **75+ yrs** | 165533 (32.92%) |
| **Race** |  |
| **White** | 369710 (73.50%) |
| **Black** | 41825 (8.31%) |
| **Asian** | 13491 (2.68%) |
| **Others** | 48988 (9.74%) |
| **Unknown** | 29011 (5.77%) |
| **Gender** |  |
| **Male** | 209292 (41.61%) |
| **Female** | 293729 (58.39%) |

**eTable 2: List of Concept Unique Identifiers (CUIs) used to identify obesity**

| **Term** | **CUI** | **Term** | **CUI** |
| --- | --- | --- | --- |
| Obesity | C0028754 | Body mass index 30+ - obesity | C0424671 |
| Morbid obesity | C0028756 | Overweight and obesity | C1561826 |
| Truncal obesity | C4551560 | Class I obesity | C4476813 |
| Pediatric Obesity | C2362324 | OBESITY AND HYPOPIGMENTATION | C5774289 |
| Simple obesity | C0451819 | Truncal obesity (recessive) | C4538693 |
| Exogenous obesity | C1281429 | Truncal obesity (variable) | C3552691 |
| Obesity, Abdominal | C0311277 | Obesity (older children) | C1862918 |
| Other obesity | C0348480 | Obesity (after infancy) | C3277018 |
| Drug-induced obesity | C0348956 | Obesity after puberty | C3277526 |
| Adolescent Obesity | C4317171 | Truncal obesity, mild | C1845451 |
| Android obesity | C0342940 | OBESITY, LATE-ONSET | C4016782 |
| Constitutional obesity | C2937224 | Obesity, early-onset | C1833056 |
| Obesity, Maternal | C0746977 | Obesity by contributing factors | C1532611 |
| Hypothyroid obesity | C0271792 | Early-onset severe obesity | C4013980 |
| Hypothalamic obesity | C0271885 | OBESITY, MILD, EARLY-ONSET | C4016893 |
| Lifelong obesity | C0267989 | Grade 3 Obesity, CTCAE | C5552857 |
| Generalized obesity | C0342942 | Grade 4 Obesity, CTCAE | C1556382 |
| Hypogonadal obesity | C0271523 | symmetrical obesity (greater than 30% over) | C2039138 |
| Infantile Obesity | C4553478 | Obesity by fat distribution pattern | C1532612 |
| Buffalo obesity | C0239194 | Obesity due to SIM1 deficiency | C5191050 |
| Hypertrophic obesity | C1260894 | Obesity due to CEP19 deficiency | C5190985 |
| Hyperinsulinar obesity | C0545039 | Obesity by adipocyte growth pattern | C1532609 |
| Peripheral obesity | C0424629 | Obesity due to pituitary disease | C5569015 |
| Gynecoid obesity | C0342941 | Obesity caused by energy imbalance | C4302408 |
| Obesity, CTCAE | C1963185 | Truncal obesity developing in midchildhood | C4538566 |
| Gross obesity | C0857116 | Obesity in females after puberty | C5543341 |
| Moderate obesity | C0694533 | Obesity, severe (onset from birth) | C5194520 |
| Obesity, hypoplasmic | C0545041 | Overweight, obesity and other hyperalimentation (E65-E68) | C1561827 |
| Obesity, hyperplasmic | C0545040 | obesity (more than 10% over ideal weight) | C2230143 |
| Sarcopenic obesity | C4063061 | Other Obesity due to Excess Calories | C2874226 |
| Hyperplastic obesity | C1532480 | OBESITY, SEVERE, AND TYPE II DIABETES | C4016768 |
| Obesity, primary | C0149974 | Onset of obesity from 6 months to 6 years | C1867874 |
| Obesity cardiomyopathy | C4552322 | Obese abdomen | C0426650 |
| Obesity, transient | C5830321 | Obese class I | C3853264 |
| Obesity, mild | C1841613 | Obese class II | C3853263 |
| centrifugal obesity | C2026325 | Type 2 diabetes mellitus in obese | C0271638 |
| secondary obesity | C2062945 | Body mass index 40+ - severely obese | C1319441 |
| Polygenic Obesity | C4054170 | Class III obesity | C1556381 |
| Iatrogenic Obesity | C4054748 | Lower body obesity | C4759305 |
| obesity; glandular | C1398735 | obesity in diabetes | C2945676 |
| pituitary; obesity | C1400190 | Adult-onset obesity | C0267990 |
| Syndromic obesity | C5680938 | Hyperplastic-hypertrophic obesity | C2937223 |
| Genetic obesity | C5681294 | Class II obesity | C1556380 |
| PECTORAL OBESITY | C0240674 | THIGH OBESITY | C0241373 |
